# Prevalence of SARS-CoV-2 in human post-mortem ocular tissues

**DOI:** 10.1101/2020.10.05.20201574

**Authors:** Onkar B. Sawant, Sneha Singh, Robert Emery Wright, Kayla M. Jones, Michael S. Titus, Eugene Dennis, Eric Hicks, Parag A. Majmudar, Ashok Kumar, Shahzad I. Mian

## Abstract

**Background:** SARS-CoV-2 is found in conjunctival swabs and tears of COVID-19 patients. However, the presence of SARS-CoV-2 has not been detected in the human eye to date. We undertook this study to analyze the prevalence of SARS-CoV-2 in human post-mortem ocular tissues.

**Methods:** The expression of SARS-CoV-2 RNA was assessed by RT-PCR in corneal and scleral tissues from 33 surgical-intended donors who were eliminated from a surgical use per Eye Bank Association of America (EBAA) donor screening guidelines or medical director review or positive COVID-19 test. Ocular levels of SARS-CoV-2 RNA (RT-PCR), Envelope and Spike proteins (immunohistochemistry) and anti-SARS-CoV-2 IgG and IgM antibodies (ELISA) in blood were evaluated in 10 COVID-19 donors.

**Findings:** Of 132 ocular tissues from 33 surgical-intended donors, the positivity rate for SARS-CoV-2 RNA was ∼13% (17/132). Of 10 COVID-19 donors, six had PCR positive post-mortem nasopharyngeal swabs whereas eight exhibited positive post-mortem anti-SARS-CoV-2 IgG levels. Among 20 eyes recovered from 10 COVID-19 donors: three conjunctival, one anterior corneal, five posterior corneal, and three vitreous swabs tested positive for SARS-CoV-2 RNA. SARS-CoV-2 spike and envelope proteins were detected in epithelial layer of the corneas that were procured without Povidone-Iodine (PVP-I) disinfection.

**Interpretations:** Our study showed a small but noteworthy prevalence of SARS-CoV-2 in ocular tissues from COVID-19 donors. These findings underscore the criticality of donor screening guidelines, post-mortem nasopharyngeal PCR testing and PVP-I disinfection protocol to eliminate any tissue harboring SARS-CoV-2 being used for corneal transplantation.

**Funding:** Research grant from EBAA and National Institutes of Health.

## Introduction

The COVID-19 pandemic caused by the Severe Acute Respiratory Syndrome Coronavirus −2 (SARS-CoV-2) has been the current focus of research as it has significantly disrupted many livelihoods. The SARS-CoV-2 virus is highly infectious and transmitted primarily through respiratory droplets and upon contact with infected persons. Studies to date have suggested that COVID-19 patients have a high viral load in the upper respiratory tract at disease onset [1, 2]. There is a strong possibility the virus may contaminate the ocular surface via respiratory droplets after coughing, sneezing, and hand-to-eye contact. In January 2020, an ophthalmologist contracted COVID-19 from an asymptomatic glaucoma patient; the initial indication was that the ocular surface served as a mediator of viral infection [3]. Studies have shown that SARS-CoV-2 may cause conjunctivitis, and viral RNA has been detected in tears and retinal biopsies of COVID-19 patients [4-11]. Conjunctival manifestations have been reported at rates as low as 0.8% [6] in the largest retrospective study from China, and at rates of 6% [9], 32% [5], and as high as 66% [10] in other studies. These reports indicate that the ocular surface may act as a possible mode of disease transmission.

SARS-CoV-2 relies on angiotensin-converting enzyme-2 (ACE-2) as its receptor on human cells, along with TMPRSS2 or Furin protease for viral entry into host cells. Ocular surface cells of the cornea and conjunctiva epithelial cells have been shown to express ACE-2 and TMPRSS2 [12, 13]. Hence, based on the recent evidence, potential SARS-CoV-2 transmission through the ocular surface remains a significant concern. The route of transmission and infiltration of the virus within the ocular tissue is still unknown. Answering these questions are of critical importance concern to ophthalmologists, eye banking industry, and the field of sight-restoring transplantation.

Although it is believed that SARS-CoV-2 primarily transmits via respiratory droplets, extra-respiratory transmission via blood is theoretically possible because studies have shown the presence of SARS-CoV-2 RNA in blood samples [14, 15]. Hence, there is a strong possibility that various ocular tissues feature different degrees of transmission risk. While few recent studies and case reports have detected viral RNA in tears and conjunctival swabs, the presence of SARS-CoV-2 has not been shown within ocular tissues of COVID-19 patients. This study was designed to systematically evaluate the presence of SARS-CoV-2 RNA and proteins in post-mortem ocular tissues of COVID-19 positive donors.

## Material and Methods

This study was performed in compliance with the Declaration of Helsinki and Eye Bank Association of America (EBAA) and Food and Drug Administration (FDA) regulations. Consent for research was obtained prior to procurement from each donor family. The University of Michigan medical school institutional review board (IRBMED) determined that this study does not fit the definition of human subjects research requiring IRB approval. Laboratory experiments from this study were approved by the Institutional Biosafety Committee (IBC) at the Wayne State University (IBC# 20-04-2164). A simplified outline of this study is provided in Fig. 1 and detailed procedures are described below.

**Figure 1:**
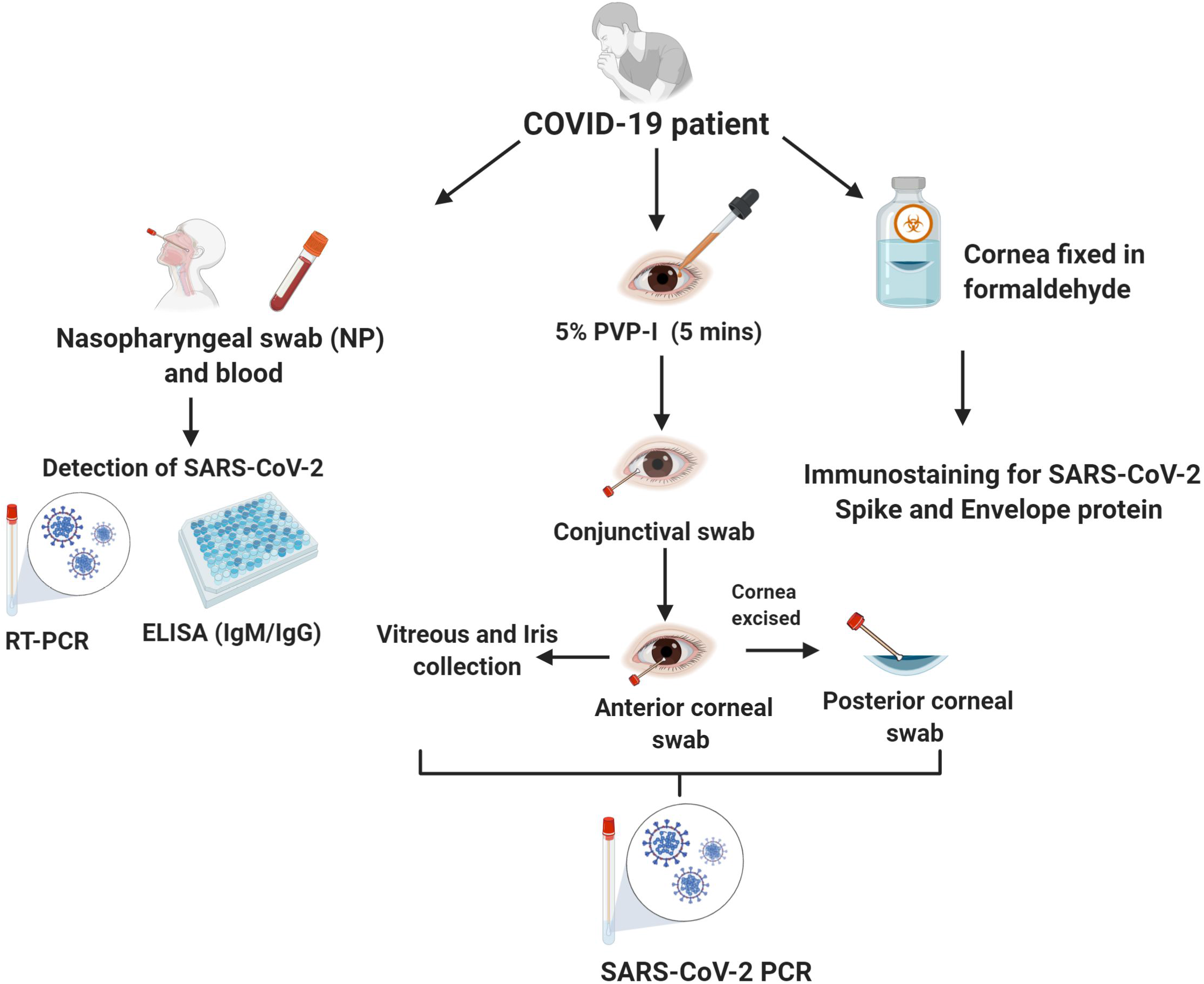
Simplified schematic representation of the procedures used for detection of SARS-CoV-2 RNA and viral antigen in various ocular tissues. Post-mortem blood and nasopharyngeal (NP) swabs were used to confirm COVID-19 followed by collection of conjunctival swab, anterior and posterior corneal swab, vitreous, and iris for RT-PCR detection of SARS-CoV-2 RNA. Fixed corneal tissues from OD (right) eyes that were procured without any PVP-I disinfection treatment were used for immunohistochemistry (IHC) detection of viral antigens.

### Donor screening criteria and procurement procedure

Donor enrollment criteria included a positive COVID-19 PCR test within 15 days of death. All 10 donors had active COVID-19 at the time of death and were reported to state agencies as COVID-19 deaths. The donors’ families had provided consent for cornea and whole eye recovery for research purposes, and all cases were in the U.S. states of Michigan or New Jersey. Appropriate personal protective equipment (PPE) and standard disposable instruments were used for eye recovery purposes. A postmortem nasopharyngeal (NP) swab and blood were collected for serological analysis. The left eye was recovered first, followed by the right eye, with disinfection of the instrument taking place between the recovery of each eye. The ocular tissue was swabbed starting with the left eye. The left eye was opened, and the anterior surface was soaked in 5% povidone-iodine (PVP-I) (Aplicare, Meriden, CT) for five minutes. The eye was thoroughly flushed with sterile eyewash solution (Medique Products, Fort Myers, FL). The lid speculum was carefully inserted, followed by a conjunctival swab collection, and the tissue was excised 360°. A sample of the conjunctival tissue was placed in a 5mL microcentrifuge tube (Eppendorf AG, Hamburg, Germany) containing 4% paraformaldehyde (PFA). The remaining tissue was soaked in PVP-I for another five minutes, followed by a sterile saline flush. A swab of the anterior cornea was taken by touching the swab to the cornea’s epithelial surface (referred to as anterior cornea). Next, the whole eye was enucleated and positioned in the RE-One chamber (RE-One Sagl, Lugano, Switzerland, acquired via Medical Innovation Partners International, Dallas, TX) for corneal excision. The cornea was removed and thereafter swabbed on the cornea’s endothelial surface (referred to as posterior cornea). The cornea was stored in a container with 4% PFA. The iris and lens were carefully removed, and the iris stored without storage media at −80 ^°C^. The vitreous was swabbed as well. The RE-One chamber was assembled, and the posterior part was submerged in a fixative solution. The instruments were disinfected using CaviWipes^™^ (Metrex, Orange, CA). The tissue recovery for each donor resulted in nine swabs for SARS-CoV-2 PCR testing (one NP, two conjunctiva, two anterior corneas, two posterior corneas, and two vitreous) and a postmortem blood sample for measuring anti-SARS-CoV-2 IgM and IgG using ELISA. The nine swabs and blood samples were immediately shipped to Eurofins VRL Laboratories (Centennial, CO USA) for SARS-CoV-2 testing by qRT-PCR.

### Diagnostic testing

RT-PCR testing was performed using The BioGX SARS-CoV-2 Reagents for BD MAX™ System at Eurofins VRL Laboratories. The U.S. Food and Drug Administration (FDA) granted Emergency Use Authorization (EUA) for The BioGX SARS-CoV-2 Reagents for BD MAX™ System on April 3, 2020. All tests were performed after that date. The BD SARS-CoV-2 Reagents for BD MAX System utilized multiplexed primers and probes targeting RNA from the nucleocapsid phosphoprotein gene (N1 and N2 regions) of the SARS-CoV-2 coronavirus, and the human RNase P gene was used as an internal control. To eliminate false positives, if the Ct value (number of cycles) was higher than 37, samples were deemed positive only if both N1 and N2 Cts were detectable with positive amplification. Otherwise, samples were deemed negative/undetectable. Serological testing for SARS-CoV-2 was performed using the EUROIMMUN Anti-SARS-CoV-2 IgG ELISA kit (Mountain Lakes, NJ USA) and the Gold Standard Diagnostics SARS-CoV-2 IgM ELISA Test Kit (Davis, CA USA) according to manufacturer’s instructions.

### RT-PCR detection for SARS-CoV-2 in ocular tissues

To assess the presence of SARS-CoV-2 in ocular tissue, we evaluated tissues that were procured for surgical purposes but later ruled out due to positive post-mortem tests or signs/symptoms of COVID-19. Corneal (n =66) and scleral (n = 66) tissue from 33 different donors were dissociated using the bead lysis method followed by total RNA extraction using Trizol reagent (Invitrogen, Carlsbad, CA) per the manufacturer’s instructions. cDNA was prepared using 1mg of RNA using a Maxima first-strand cDNA synthesis kit (Thermo Scientific, Waltham, MA) per the manufacturer’s instructions. Viral RNA was detected in the cDNA using SARS-CoV-2 nucleoprotein gene-specific Taqman probe primers purchased from Integrated DNA Technologies (Coralville, IA). The primer sequence used in the assay was adopted from the Chinese Center for Disease Control and Prevention primers for detection of 2019-nCoV: Forward primer: GGGGAACTTCTCCTGCTAGAAT; Reverse primer: CAGACATTTTGCTCTCAAGCTG; Fluorescent probe: 5’-FAM-TTGCTGCTGCTTGACAGATT-TAMRA-3’. RT-PCR was conducted using the StepOnePlus Real-Time PCR system (Applied Biosystems, Foster City, CA). The quantification of gene expression was determined by the Standard Quantification method and expressed as Ct values. Tissue samples with Ct value ≤37 were considered positive, while undetermined Ct values and Ct values >37 were considered negative. The whole-genome SARS-CoV-2 RNA was used as a positive control, while a no-template mixture was used as a negative control for the assay.

### Immunohistochemistry

Fixed corneas were passed through a series of sucrose gradient (10%, 20%, 30%), and embedded in OCT (Tissue-Tek^®^-Sakura, Torrance, CA). Ten-micrometer thin sections were prepared using a cryotome (HM525 NX, ThermoFisher Scientific, Waltham, MA) and mounted onto lysine-coated glass slides (Fisherbrand, Thermo Scientific, Waltham, MA). The sections were permeabilized and blocked with 10% normal goat serum with 0.5% Triton X-100 for two hours at RT and incubated overnight with primary mouse anti-SARS-CoV-2 spike or envelope antibodies (BEI Resources, NIAID) (1:100). The next day, sections were rinsed four times with PBS (10 minutes each) and incubated with anti-mouse/rabbit Alexa Fluor 485/594-conjugated secondary antibodies (1:200) for 2 h at RT. The sections were extensively rinsed with PBS (four washes, 10 minutes each) and the slides were mounted in Vectashield anti-fade mounting medium (Vector Laboratories, Burlingame, CA) and visualized using a Keyence microscope (Keyence, Itasca, IL) at different magnifications.

## Results

### PCR testing on surgical rule-out tissues

To detect the presence of SARS-CoV-2 RNA, we isolated RNA from the sclera and cornea (without separating different layers) of asymptomatic positive donors and donors that had symptoms of COVID-19 without positive COVID-19 PCR test. All of these donors were classified in one of three different groups, and their testing results are summarized in Table 1. Group 1 primarily consisted of eye-only donors who had passed EBAA screening criteria with donors that were deemed appropriate for surgical use. We performed NP swab at the time of corneal recovery and received a positive test result. Group 2 primarily consisted of donors early in the pandemic when testing was not widespread. Corneas were ruled out for surgical use after consultation with medical directors or following the application of EBAA donor eligibility guidelines based on signs and symptoms. The majority of these cases had a negative COVID-19 PCR test. Group 3 consisted of donors who neither had signs or symptoms nor tested positive for COVID-19 but had spent a significant amount of time with someone with COVID-19. Results from SARS-CoV-2 RT-PCR testing are summarized in Table 1. The highest positivity rate of 17% for sclera and 11% for cornea was observed in Group 1. In Group 2, 12% of scleral samples and 15% of corneal samples were positive for SARS-CoV-2 viral RNA. None of the tissues from the two donors in Group 3, who had close contact with a COVID-19 patient, showed any detectable level of SARS-CoV-2 RNA.

**Table 1:** SARS-CoV-2 PCR results for surgically ruled-out tissues

| **Groups** | **COVID-19 Positive**<br>**(Group 1)** |  | **Signs and Symptoms of**<br>**COVID-19**<br>**(Group 2)** |  | **Close Contact With**<br>**COVID-19 Patient**<br>**(Group 3)** |  |
| --- | --- | --- | --- | --- | --- | --- |
| **Number of Cases** | 18 |  | 13 |  | 2 |  |
| **Median Age (Range)** | 61 (17-72) |  | 59 (26-75) |  | 63 (62-63) |  |
| **Tissue Type** | Sclera | Cornea | Sclera | Cornea | Sclera | Cornea |
| **Number of Tissues** | 36 | 36 | 26 | 26 | 4 | 4 |
| **RNA Concentration**<br>**(Ng/μl)** | 520 ± 61 | 537 ± 66 | 539 ± 50 | 486 ± 41 | 591 ± 65 | 522 ± 69 |
| **Number of Positive**<br>**Tissues** | 6 | 4 | 3 | 4 | 0 | 0 |
| **\*Ct (Mean ± SD)** | 33 ± 2 | 32 ± 2 | 35 ± 1 | 29 ± 7 |  |  |
| **Positivity Rate** | **17%** | **11%** | **12%** | **15%** | **0%** | **0%** |

\*Ct refers to number of PCR cycles

### Detecting SARS-CoV-2 in donors that died due to COVID-19

After observing 13% (17 out of 132) SARS-CoV-2 RNA prevalence rate in surgical rule-out tissues, our next goal was to systematically evaluate the presence of SARS-CoV-2 in different ocular layers of research consented donors that died due to COVID-19. We analyzed ocular swabs from 10 different donors with a median age of 66 (range 46-90). Donor demographics and pre-mortem testing results are provided in Table 2. Detailed case summaries with a timeline of disease progression and testing results are provided below. The testing results are also summarized in Table 3.

**Table 2:** Donor demographics and pre-mortem testing results

| **Case #** | **Gender** | **Race** | **Pre-Mortem COVID-19**<br>**Test Result** | **Pre-Mortem Testing to Death**<br>**Interval (days)** |
| --- | --- | --- | --- | --- |
| 1 | Male | Caucasian | Positive | 6 |
| 2 | Female | Caucasian | Positive | 9 |
| 3 | Male | Caucasian | Positive | 7 |
| 4 | Male | South Asian | Positive | 15 |
| 5 | Female | Caucasian | Positive | 3 |
| 6 | Male | Hispanic | Positive | 10 |
| 7 | Female | Caucasian | Positive | 3 |
| 8 | Female | Hispanic | Positive | 11 |
| 9 | Male | Black | Positive | 1 |
| 10 | Male | Hispanic | Positive | 3 |

**Table 3:** Post-mortem SARS-CoV-2 testing results

| Case # | NP-PCR | IgG | IgM | Eye | Conjunctiva | Anterior Cornea | Posterior Cornea | Vitreous |
| --- | --- | --- | --- | --- | --- | --- | --- | --- |
| Case #1 | Positive | Positive | -ve | Right | -ve | -ve | Positive | -ve |
|  |  |  |  | Left | -ve | -ve | -ve | -ve |
| Case #2 | Positive | -ve | -ve | Right | -ve | -ve | -ve | -ve |
|  |  |  |  | Left | -ve | -ve | Positive | -ve |
| Case #3 | -ve | Positive | -ve | Right | -ve | -ve | -ve | -ve |
|  |  |  |  | Left | -ve | -ve | -ve | -ve |
| Case #4 | -ve | Positive | -ve | Right | -ve | -ve | -ve | -ve |
|  |  |  |  | Left | -ve | -ve | -ve | -ve |
| Case #5 | Positive | Positive | -ve | Right | -ve | -ve | -ve | -ve |
|  |  |  |  | Left | -ve | -ve | -ve | -ve |
| Case #6 | Positive | Positive | Positive | Right | -ve | -ve | -ve | -ve |
|  |  |  |  | Left | Positive | -ve | -ve | -ve |
| Case #7 | -ve | Positive | -ve | Right | -ve | -ve | Positive | -ve |
|  |  |  |  | Left | -ve | -ve | -ve | Positive |
| Case #8 | -ve | Positive | -ve | Right | -ve | -ve | Positive | -ve |
|  |  |  |  | Left | -ve | -ve | -ve | -ve |
| Case #9 | Positive | -ve | -ve | Right | Positive | Positive | Positive | Positive |
|  |  |  |  | Left | Positive | -ve | -ve | Positive |
| Case #10 | Positive | Positive | -ve | Right | -ve | -ve | -ve | -ve |
|  |  |  |  | Left | -ve | -ve | -ve | -ve |
| **Positivity Rate** | **60%** | **80%** | **10%** |  | **15%** | **5%** | **25%** | **15%** |

#### Case Summaries

**Case #1:** A Caucasian male in his 60s was admitted for potentially infected lower extremity ulcers and type I diabetes mellitus (DM). On the same day, the patient tested positive for COVID-19 by PCR. Follow-up positive COVID-19 PCR tests were reported 13 and 28 days after hospital admission. The patient passed away six days after last positive COVID-19 test. Post-mortem NP swab test was positive for SARS-CoV-2 RNA along with the presence of anti-SARS-CoV-2 IgG-Ab. Among the ocular tissue swabs, the right posterior cornea tested positive for SARS-CoV-2 RNA. All other ocular tissue swabs were negative.

**Case #2:** A Caucasian female in her late 80s was presented to an emergency room (ER) with fever, worsening cough, shortness of breath, body aches, and poor oral intake. On the same day, the patient tested positive for COVID-19 via PCR assay. The patient subsequently passed away nine days after testing positive for COVID-19. The post-mortem NP swab test was positive for SARS-CoV-2 RNA while the anti-SARS-CoV-2 IgG Ab test was negative. Among the ocular tissue swabs, the left posterior cornea tested positive for SARS-CoV-2 RNA. All other ocular tissue swabs were negative.

**Case #3**: A Caucasian male in his early 70s was admitted to the hospital with symptoms of respiratory distress. On the same day, the patient was tested positive for COVID-19 via PCR assay. The patient passed away seven days after hospital admission and testing positive for COVID-19. Surprisingly, the post-mortem NP swab test was negative for SARS-CoV-2 RNA. However, the anti-SARS-CoV-2 IgG-Ab test was positive indicating that this donor might have had SARS-CoV-2 infection well before admission to the hospital. All ocular tissue swabs were negative.

**Case #4:** A South Asian male in his 60s was admitted to the hospital with right middle cerebral artery stroke. The patient tested positive for COVID-19 via PCR assay after two months of hospital stay. The patient subsequently passed away 15 days after testing positive for COVID-19. Similar to case #3, the post-mortem NP swab test was negative for SARS-CoV-2 RNA. However, the anti-SARS-CoV-2 IgG Ab test was positive, indicating that longer duration between pre-mortem testing and time of death may not yield positive post-mortem tests due to a reduction in viral load in the upper respiratory tract. All ocular tissue swabs were negative for viral RNA.

**Case #5:** A Caucasian female in her 70s was admitted to the hospital with symptoms of respiratory distress. The patient tested positive for COVID-19 by PCR assay on the same day and 23 days after hospital admission. The patient passed away three days after final positive COVID-19 test. The post-mortem NP swab test was positive for SARS-CoV-2 RNA, and the anti-SARS-CoV-2 IgG Ab test was positive as well, indicating prolonged active infection with seroconversion. However, to our surprise, all ocular tissue swabs were negative.

**Case #6:** A Hispanic male in his late 60s was admitted to the hospital after being found unresponsive. The patient was deemed positive for COVID-19 via PCR assay on the next day after hospital admission. The patient passed away 10 days after testing positive for COVID-19. The post-mortem NP swab test for the presence of SARS-CoV-2 RNA, anti-SARS-CoV-2 IgG, and IgM Abs were positives. Among the ocular tissue swabs, the left conjunctiva tested positive for SARS-CoV-2 RNA. All other ocular tissue swabs were negative.

**Case #7:** A Caucasian female in her late 50s was admitted to the hospital with a fever of 102.8°F, progressive weakness over the past few days, and a dry cough for the past two weeks. On the same day, the patient tested positive for COVID-19 via PCR assay and tested positive again on 12^th^ day after hospital admission. The patient subsequently passed away three days after final positive COVID-19 test. This was the first case we assessed in which ocular tissues tested positive for SARS-CoV-2 RNA while the post-mortem NP swab was negative for viral RNA. Among the ocular tissue swabs, the right posterior cornea and left vitreous tested positive. All other ocular tissue swabs were negative. This donor also had positive IgG antibodies against SARS-CoV-2.

**Case #8:** A Hispanic female in her early 90s was admitted to the hospital and tested positive for COVID-19 by PCR assay on the same day. The patient passed away 11 days after testing positive for COVID-19. Similar to Case #4, the post-mortem NP swab test was negative but the anti-SARS-CoV-2 IgG Ab test was positive, probably due to a relatively longer duration between pre-mortem testing and time of death. Interestingly, among the ocular tissue swabs, the right posterior cornea tested positive for SARS-CoV-2. All other ocular tissue swabs were negative.

**Case #9:** An African American male in his late 40s was admitted to the hospital for difficulty in breathing, productive cough, abdominal distension, and worsening lower extremity edema. The patient tested positive for COVID-19 by PCR assay on the same day. He passed away next day after hospital admission. Post-mortem NP swab testing for the presence of SARS-CoV-2 RNA was positive but the anti-SARS-CoV2-IgG Ab test was negative, indicating early-stage infection with incomplete seroconversion. This donor had positive swabs from both conjunctivae, right anterior cornea, right posterior cornea, and both vitreous. Left anterior and posterior corneal swabs tested negative.

**Case #10:** A Hispanic male in his 40s was admitted to the hospital for flu-like symptoms that developed six days before admission. He tested positive for SARS-CoV-2 via PCR tests that were performed on the day of admission and 42 days after admission. The patient passed away 3 days after final positive COVID-19 test. Similar to Case #3, the post-mortem NP swab test was positive for SARS-CoV-2 RNA and anti-SARS-CoV-2-IgG Ab test was positive, indicating prolonged active infection along with seroconversion. However, all ocular tissue swabs were negative indicating that SARS-CoV-2 RNA might be absent from ocular tissues despite positive post-mortem NP swab if the patient had COVID-19 for a longer duration.

In summary, of the 10 donors, we determined six donors had positive post-mortem NP swabs and 8 donors had positive anti-SARS-CoV-2-IgG antibodies. Two donors (cases 2 and 9) with negative IgG exhibited positive NP swabs, which might indicate an early stage of infection without seroconversion. Nonetheless, all 10 donors had either a positive post-mortem NP swab or a positive IgG, validating pre-mortem testing, and diagnosis of COVID-19. Across all 20 eyes: Three conjunctival, one anterior corneal, five posterior corneal, and three vitreous swabs tested positive for SARS-CoV-2 RNA, exhibiting a positivity rate of 15% for conjunctiva, 5% for anterior corneal surface, 25% for posterior corneal surface, and 15% for vitreous (Table 3).

### Immunohistochemical Validation

Right corneal tissue that were procured without any PVP-I disinfection and showed overall stronger positivity (lower CT value) for SARS-CoV-2 was immunostained to detect viral antigens. Our IHC data showed that coinciding with positive viral RNA, SARS CoV-2 Envelope protein (Fig. 2), and Spike protein (Fig. 3) were detected in the COVID-19 patient’s right corneas (Cases 7-9). The positive staining was primarily observed in epithelial layer of the cornea. In contrast, no positivity for spike protein was visualized in COVID-19 negative patient corneas (healthy control). Non-specific antibody binding was ruled out by staining sections with a secondary antibody only as control. Also, the specificity of antibodies was validated by infecting cells with live SARS-CoV-2 and staining them for spike protein (data not shown).

**Figure 2:**
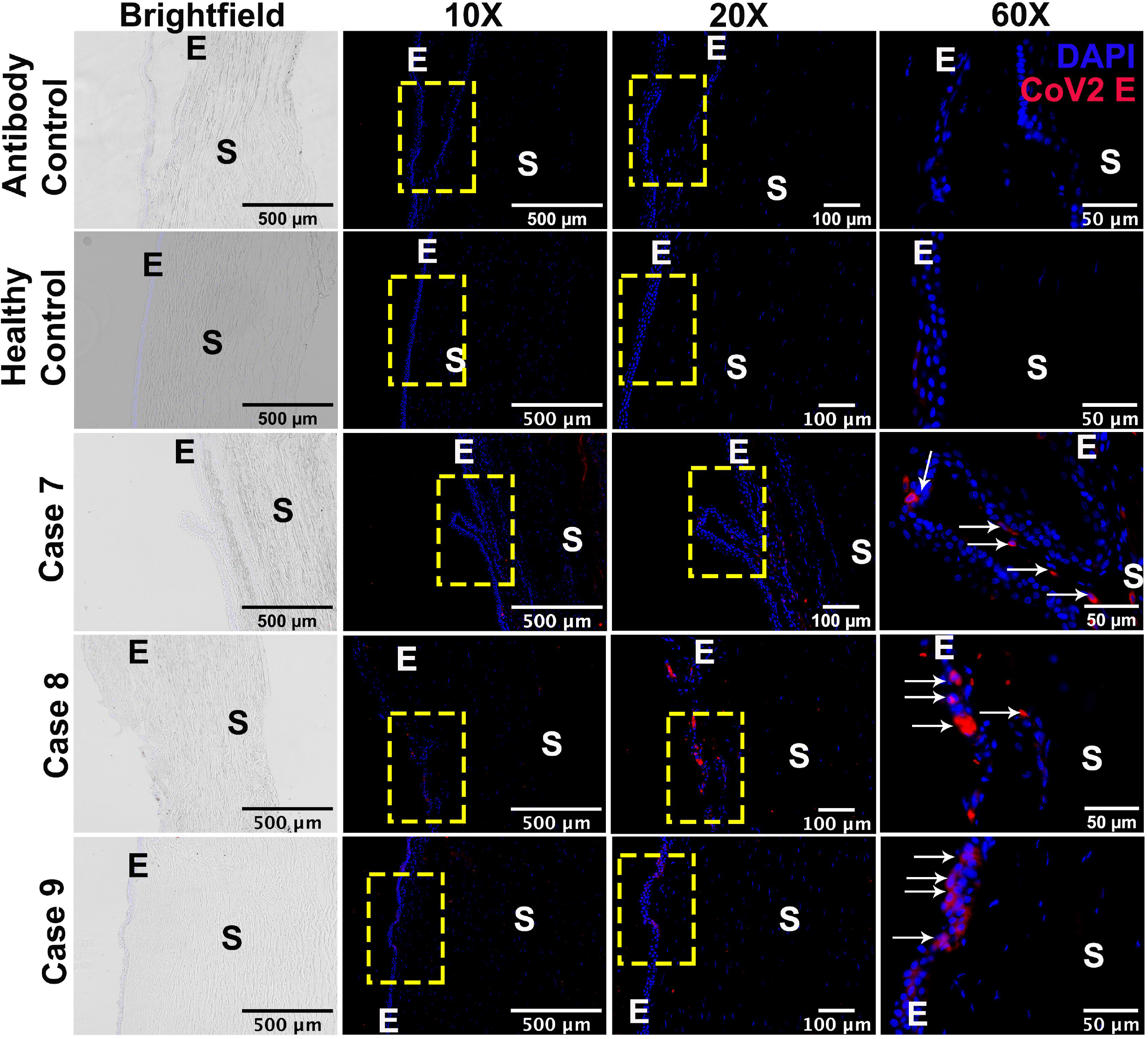
SARS-CoV-2 Envelope (E) protein was detected in the cornea of the COVID-19 donors that were procured without any PVP-I disinfection treatment. OD (right) cornea from healthy and COVID-19 donors were fixed in formaldehyde and 10μm thin sections were stained for IHC using antibody against SARS-CoV-2 Envelope (E) protein (red color) while DAPI was used for nuclear staining (blue color). The image was captured at different magnifications (10X, 20X, and 60X) to visualize cellular location of the viral proteins. The region of interest has been highlighted using a yellow box and white arrows. **E**, corneal epithelium; **S**, corneal stroma. Sections stained with secondary antibody (anti-mouse Alexa Fluor 594) was used to assess the antibody specificity.

**Figure 3:**
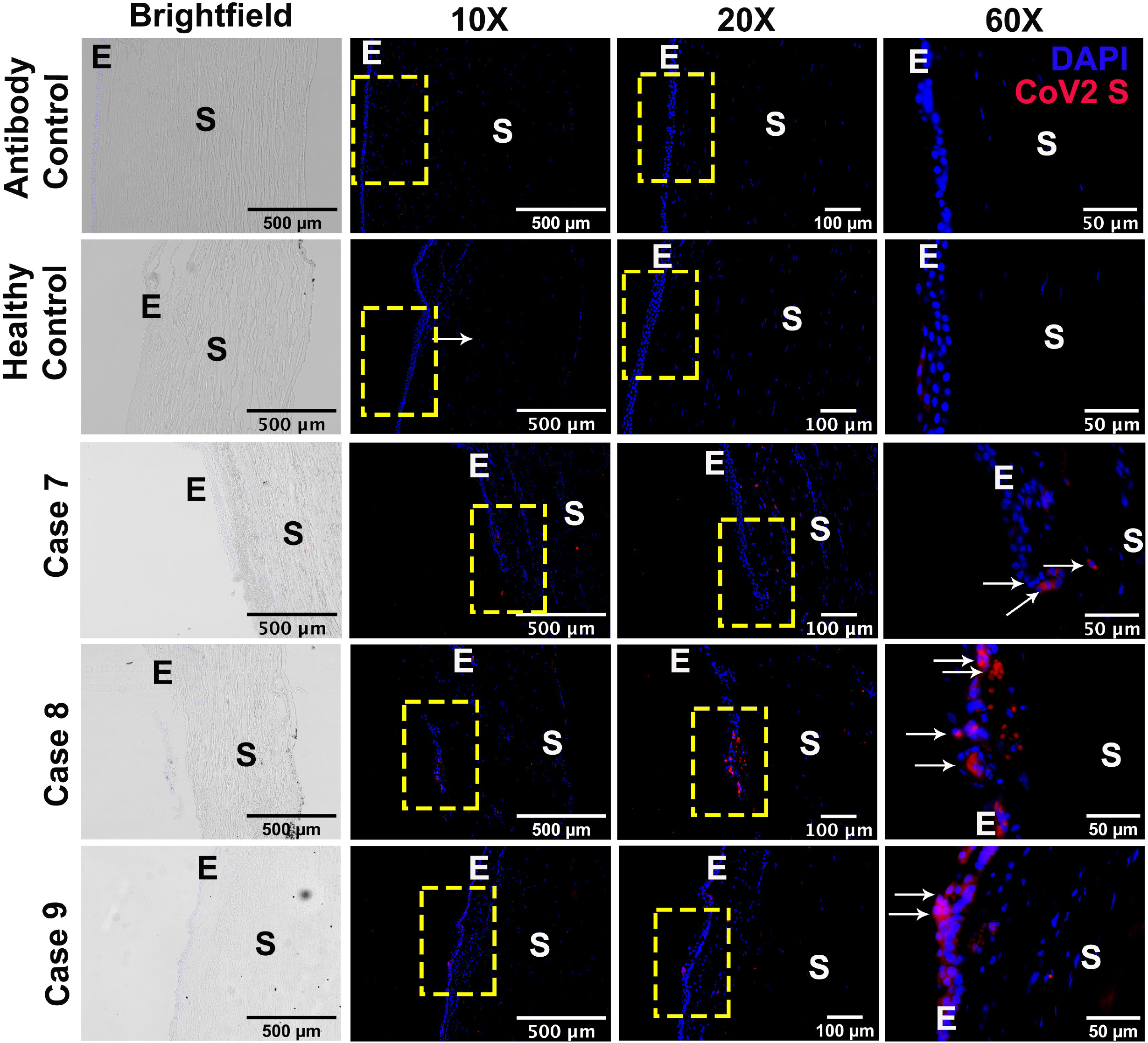
SARS-CoV-2 Spike (S) protein was detected in the corneal epithelium of the COVID-19 donors that were procured without any PVP-I disinfection treatment. OD (right) cornea from healthy and COVID-19 donors were fixed in formaldehyde and 10μm thin sections were stained for IHC using antibody against SARS-CoV-2 Spike (S) protein (red color) while DAPI was used for nuclear staining (blue color). The image was captured at different magnifications (10X, 20X, and 60X) to visualize cellular location of the viral proteins. The region of interest has been highlighted using a yellow box and white arrows. **E**, corneal epithelium; **S**, corneal stroma. Sections stained with secondary antibody (anti-mouse Alexa Fluor 594) was used to assess the antibody specificity.

## Discussion

Cornea transplant is usually a safe ophthalmic procedure with little morbidity. However, there is always a potential risk for transmission of infectious agents, especially viruses, from donors to recipients or causing graft complications [16]. Eye banks follow elaborate procedures to mitigate this risk through extensive donor screening including testing for the presence of microbial pathogens (REF). Because of increased mortality due to COVID-19 and high transmissibility of the SARS-CoV-2 virus, we investigated its prevalence in ocular tissues, which is currently unknown. In this study, for the first time, we report the presence of not only SARS-CoV-2 RNA but also its envelope and spike proteins in ocular tissues of donors who had COVID-19 at the time of death. SARS-CoV-2 RNA was also detected in 13% (17 out of 132) corneal and scleral tissues recovered from 33 asymptomatic positive carriers or symptomatic negative donors. Overall, our findings underscore the criticality of post-mortem PCR testing, PVP-I disinfection protocol and donor screening guidelines in eye banking to eliminate the possibility of handling tissues with SARS-CoV-2 RNA particles. It is important to note that, while our data showed the presences of viral RNA and antigens in the corneas of COVID-19 patients, further studies are necessary to explore the possibility of transmission via ocular tissues and concentration dependent anti-virulent activity of PVP-I.

In the context of the route of administration, it has been proposed that the virus travels via direct airborne transmission since the entire ocular surface including the anterior cornea, tear film and conjunctiva communicates with air, similar to the nasal mucosa. Another proposed path of infiltration is the nasolacrimal route, facilitating the transport of viral particles from the respiratory tract to the ocular surface or vice versa [17]. These potential routes of infiltration may justify reports suggesting that SARS-CoV-2 causes conjunctivitis in COVID-19 positive patients [4-6, 9, 10, 18]. Incidences of conjunctivitis in these reports are mainly in the range of 0.8-6% but they can be as high as 32% and 66% (for a detailed review, please refer to [19]).

Although the primary route of SARS-CoV-2 infiltration could be via respiratory droplets, a potential non-respiratory route of transmission cannot be ignored. To our surprise, we observed a higher prevalence of SARS-CoV-2 RNA in the posterior corneal (endothelial) surface than from any other ocular surface swabs (Table 3). This finding and current literature highlighting the prevalence of SARS-CoV-2 in extra-respiratory routes such as blood [14, 15] led us to hypothesize that vascular endothelium-enriched ocular tissues could be the source of posterior corneal infectivity. To test this hypothesis, we analyzed iris samples from COVID-19 donors for the presence of SARS-CoV-2 RNA. Surprisingly, none of the iris samples showed detectable levels of SARS-CoV-2 RNA via PCR test (data not shown). We also detected SARS-CoV-2 RNA in two of 20 vitreous swabs. Similarly, a recent study demonstrated 21% (3/14) prevalence rate of SARS-CoV-2 RNA in the retinal biopsies of deceased COVID-19 patients [11]. Hence, the possibility of infiltration via retinal vasculature could not be eliminated. Our findings of a higher prevalence of SARS-CoV-2 RNA in the posterior corneal (endothelial) surface than on any other ocular surface remain intriguing, and further studies are required to decipher the detailed mechanism. As postulated by Ang and colleagues [20], a contributing factor for this finding could be higher antimicrobial activity on the ocular surface. Tear film contains antimicrobial protein lactoferrin, which has been postulated to prevent bindings of SARS-CoV-2 to the ACE2 receptors on anterior corneal and conjunctival surfaces.

An initial study goal was to detect the effectiveness of PVP-I in inactivating the SARS-CoV-2. Therefore, we recovered right eyes without PVP-I exposure, and left eyes were recovered following EBAA-recommended double PVP-I soak procedure. We are unable to make conclusive remarks about the effectiveness of PVP-I, due to the small sample size (10 cases) of this study. However, we note that all of the anterior corneal surface swabs from left eyes (PVP-I treated) were negative for SARS-CoV-2 RNA, whereas one out of 10 right eye swabs was positive. Although many studies have demonstrated the virucidal activity of PVP-I against other similar viruses such as MERS-CoV and SARS-CoV [21-24], there is no literature explaining the mechanism of action of PVP-I to inactive SARS-CoV-2. The possibility of PVP-I mediated SARS-CoV-2 RNA degradation could not be eliminated. There is still a possibility that the PCR testing amplified non-intact SARS-CoV-2 RNA. As we learn more about the genomic sequence of the virus, more efforts should be made toward designing better PCR primers that are indicative of RNA cleavage patterns and resistant to mutational changes in the SARS-CoV-2. Such specific primers will help us classify ineffective positive PCR testing from a non-infective positive PCR test.

Respiratory viruses, including influenza virus have shown tropism towards the eye [25, 26] and have been reported to infect and replicate at the ocular surface [27]. To our knowledge, our study is the first, to show SARS-CoV-2 antigens in ocular tissue of COVID-19 patients that were procured without PVP-I disinfection treatment, supporting the idea that SARS-CoV-2 could infect corneal epithelial cells. We previously showed that corneal epithelia cells elicit innate responses upon viral stimuli such as ZIKV [28] or PolyI:C, a viral mimic of dsRNA [29]. However, further studies are necessary to understand if potential transmission of SARS-CoV-2 through the ocular surface is possible.

In summary, we report the presence of SARS-CoV-2 RNA and proteins in the ocular tissues of donors who had COVID-19 at the time of death or had related signs and symptoms. It is unclear whether the presence of SARS-CoV-2 RNA and proteins is due to primary ocular surface infection or due to retrograde transport of viral particles from the upper respiratory tract via the nasolacrimal duct. It is also unclear whether SARS-CoV-2 can replicate in corneal and/or conjunctival cells and what changes occur in infected ocular surface cells. Further studies including live virus culture, the infectivity of ocular surface cells, and elucidation of biomarkers for viral infections such as induction of inflammatory and antiviral responses are needed to establish the link that the eye is a portal for entry and transmission of SARS-CoV-2. Nonetheless, our study strongly recommends simultaneous implementation of post-mortem PCR testing, PVP-I disinfection protocol and thorough donor screening according to EBAA and CDC guidelines to mitigate the potential risk of transplanting a tissue with SARS-CoV-2 particles.

## Data Availability

We (investigators) have access to all the data represented in this manuscript.

## Acknowledgments

This work was supported by research grants from the Eye Bank Association of America (SIM, OBS, MST, PAM, AK), National Eye Institute (AK – EY027381, EY026964), and National Institute of Allergy and Infectious Diseases (AK – AI135583, AI140033). We also acknowledge our organ procurement organization (OPO) partners Gift of Life Michigan and New Jersey Sharing Network for referring COVID-19 donors for this study and Eversight’s Donation Support Center (DSC) for screening these donors. We thank Miracles in Sight (Winston-Salem, NC) and Transplant Services Center-UT Southwestern Medical Center (Dallas, TX) for contributing donated tissues to this study. Last and importantly, we thank the donors and their families whose gifts of donated eye tissue and consent for research is helping us advance the science and understanding of COVID-19 to benefit all those in need of safe transplantation and the gift of sight.

